# Genomic Epidemiology of CHIKV During the Largest Outbreak in Mainland China-Implication for CHIKV Intervention

**DOI:** 10.64898/2026.06.16.26355213

**Authors:** Lina Yi, Silin Xiang, Xuhe Huang, Jianhua Huang, Mingji Chen, Heshi Long, Yuwei He, Chuting Zeng, Guanghu Zhu, Shuyan Tan, Zhe Liu, Shanshan Gao, Jing Lu

**Author notes:** These authors contributed equally to this work (Lina Yi, Silin Xiang, Xuhe Huang, Jianhua Huang). Corresponding author. E-mail address (Jing Lu).

## Abstract

Chikungunya virus (CHIKV) has caused recurrent epidemics across tropical and subtropical regions globally. In 2025, Guangdong, China, experienced the largest documented CHIKV outbreak in mainland China, with 23,464 reported cases across all 21 prefecture-level cities. We conducted integrated epidemiological, genomic, and phylodynamic analyses to investigate the outbreak’s origins, transmission dynamics, and CHIKV lineages evolution. The Guangdong strain belongs to the East/Central/South African (ECSA) genotype, Middle African lineage (MAL; ECSA-MAL) — one of several CHIKV lineages that co-circulated globally in 2025; a long internal branch separates it from previously sequenced relatives, indicating substantial surveillance gaps in endemic regions. Phylodynamic analysis using a local molecular clock model estimated the viral introduction occurred within a narrow window in early April 2025 (95% HPD: March 26-April 12) suggesting a 2.5-month cryptic transmission prior to detection. The prolonged silent phase aligns with vector dynamics: the Mosquito Ovitrap Index (MOI) in the outbreak town of Beijiao, Foshan, stayed low in April (3.0) before surging to 9.0 through July—consistent with suppressed early transmission followed by seasonal amplification. Human migration from two epicenters (Foshan and Jiangmen) explained ∼50% of case distribution variance, while mosquito ecology constrained spatial spread. Phylowave analysis on global circulating strains identified 33 lineage-defining mutations across 14 epidemic sublineages, concentrated in NSP3, E1 and E2. Four of these have been experimentally validated as adaptive mutations. This study demonstrates the critical need for enhanced genomic surveillance during pre-peak months and underscores the importance of monitoring adaptive mutations across diverse ecological regions.

## 1 Introduction

Chikungunya fever (CHIKF) is an acute febrile illness characterized by severe polyarthralgia, myalgia, headache, and rash, often progressing to chronic arthritic symptoms persisting for months to years [1]. The causative agent, chikungunya virus (CHIKV), is an enveloped positive-sense single-stranded RNA virus belonging to the *Alphavirus* genus (family *Togaviridae*). Since its initial isolation in Tanzania in 1952–1953, CHIKV has caused numerous epidemics across Africa, Asia, Europe, and the Americas, with an estimated 1.4 billion people living in at-risk regions globally [2]. The virus is primarily transmitted by *Aedes aegypti* and *Aedes albopictus* mosquitoes, though sylvatic cycles involving non-human primates and forest-dwelling *Aedes* species maintain enzootic circulation in Africa [1].

CHIKV exhibits substantial genetic diversity and could be classified into three major genotypes: West African (WA), East/Central/South African (ECSA), Asian Urban (AUL) [3]. The ECSA genotype is further subdivided into ECSA-SAL (South American Lineage), ECSA-IOL (Indian Ocean Lineage), and ECSA-MAL (Middle African Lineage) [4]. The global distribution of CHIKV genotypes has shifted dramatically over the past two decades. The 2004–2010 Indian Ocean epidemic (IOL) and the 2013-2014 Americas invasion (ECSA-SAL) represent major invasion events facilitated by adaptive evolution and human mobility. Climate change, expanding urbanization, and increasing international travel have created new ecological niches for *Aedes* mosquitoes, potentially enabling CHIKV establishment in previously non-endemic regions. Southern China, particularly Guangdong Province, provides suitable ecological conditions for CHIKV transmission due to its subtropical climate and established *Ae. albopictus* populations. *Ae. albopictus* is the dominant *Aedes* species in Guangdong [5], with peak abundance during summer months (June–August) [6].

The year 2025 was marked by the concurrent circulation of multiple CHIKV lineages across continents. Between 1 January and 30 September 2025, a total of 445,271 suspected and confirmed CHIKV infections and 155 deaths were reported worldwide [7]. In the Americas, the ECSA-SAL lineage, drove large epidemics, with approximately 228,591 cases reported through mid-August 2025, over 95% attributable to Brazil. In South Asia, the ECSA-IOL lineage persisted in India, 30,876 suspected cases within the first quarter of 2025 alone. In the Indian Ocean, the ECSA-MAL lineage re-emerged on Réunion Island in August 2024, causing the territory’s largest epidemic since 2005–2006, with over 54,000 laboratory-confirmed cases and 40 deaths by September 2025, and spread to Mayotte in March 2025 [8]. Historically, CHIKV activity in mainland China has been limited. Small-scale outbreaks were documented without large-scale sustained transmission [9,10]. In 2025, Guangdong Province experienced the largest CHIKV outbreak in mainland China’s history [11]. In this study, we conducted an integrated epidemiological-genomic analysis to investigate the spatiotemporal dynamics of the 2025 chikungunya virus (CHIKV) outbreak in Guangdong. We illustrated the dynamics of vector density and population flow, exploring their relationships to viral transmission. Additionally, we performed Bayesian phylodynamic and lineage fitness analyses to estimate the viral introduction time and identify adaptive mutations potentially relevant to lineages transmission in different ecological regions. This work represents the comprehensive molecular epidemiological characterization of CHIKV evolution, providing critical insights for future surveillance and control in mainland China and also other risk regions.

## 2 Materials and Methods

### 2.1 Sample Collection and Data Integration

Acute-phase serum samples were collected under ethical approval granted by the Institutional Review Board of the Guangdong Provincial Institute of Public Health (IEC2025006), and informed consent was obtained from all participants [12].

*Aedes albopictus* surveillance data were obtained from the Guangdong Provincial Vector Surveillance System, which monitors all 1,656 townships/subdistricts monthly (ovitrap) and twice monthly (Breteau Index). Records from Foshan during April 1 to July 31, 2025 were aggregated by district and calendar month. Two indices were analyzed: the Mosquito Ovitrap Index (MOI; positive ovitraps per 100 ovitraps deployed) and the Breteau Index (BI; positive containers per 100 households surveyed).

Inter-city population mobility data (July–December 2025) were acquired from the Baidu Migration (Qianxi) platform (https://qianxi.baidu.com), to assess correlations between population outflow from outbreak epicenters and secondary case counts. For each of the 21 prefecture-level cities, daily outflow percentages from July 1 to December 31, 2025 were retrieved; the migration flow percentage for an origin–destination pair is the proportion of the origin city’s total daily outflow directed to that destination. To account for temporal variation, we calculated the average daily outflow percentages across two distinct phases: Phase 1 (July–August 2025), during which Foshan (Shunde District) served as the primary epicenter, and Phase 2 (September–December 2025), during which Jiangmen emerged as the dominant epicenter following the biphasic epidemic pattern. Spearman rank correlation coefficients (ρ) were used to assess the monotonic association between population mobility flow and cumulative CHIKV case counts across the 20 non-epicenter cities, given the non-normal distribution of both variables. Statistical significance was set at α = 0.05. Log-log linear regression was applied to model the power-law relationship between migration flow and case counts.

### 2.2 Sequencing, Dataset and Phylogenetic Analysis

All clinical specimens were handled, and viral RNA extraction were performed, under BSL-2 conditions in the biosafety laboratories of the Guangdong Provincial Center for Disease Control and Prevention, in accordance with national biosafety regulations. CHIKV RNA was detected with real-time RT-PCR kit (DA1911, DaAn Gene). Samples with a cycle threshold (Ct) value ≤ 35 were considered eligible for whole-genome sequencing. Multiplex RT-PCR was performed, and the generated products were sequenced on an Illumina MiSeq platform as previously described [13]. Consensus genomes were assembled by mapping reads to the S27 prototype reference (accession number AF369024) using minimap2 [14] and bcftools [15]. Per-sample sequencing results—Ct value, raw data volume, breadth of genome coverage, mean sequencing depth, and genome completeness—are provided in Supplementary Table S1. CHIKV genomes were downloaded from NCBI GenBank (n = 3,422) and GenBase (n = 106) on 7 February 2026 and combined with the genomes newly generated in this study (3,567 genomes in total). Following quality control—requiring >80% genome coverage, <10% ambiguous sites, complete country information, and sampling dates specified at least to the year—we constructed three datasets: (1) the full dataset (n = 2,486, spanning 1952–2025); (2) the last two decays dataset (sequences collected from the 2005-onwards; n = 2,410); (3) the Guangdong outbreak sequences combined with related ECSA-MAL lineage genomes (n = 247; 159 Guangdong 2025 genomes, 83 Réunion 2024–2025 genomes, and 5 sister-branch sequences). Because pre-2005 sampling is sparse (n = 52, 1952–2004) and provides no useful calibration for the recent radiation while biasing root placement, the 2005-onwards phylogeny is presented for clarity and compactness, and for following phylogenetic analyses. For dataset (3), complete year–month–day collection dates were additionally required: because the precision of tip-dated TMRCA estimation depends directly on the temporal resolution of the sampling dates. Sequences were aligned via MAFFT version 7 [16]. We inferred a maximum-likelihood phylogeny using IQ-TREE2 [17] and constructed a time-scaled tree using TreeTime (via augur refine v30.0.0) [18]. The joint date inference, an optimized coalescent model, mutation-count divergence units, and TreeTime’s default root determination (best root from root-to-tip regression) were applied.

### 2.3 Bayesian Phylodynamics

To estimate the outbreak’s introduction time, a localized Bayesian phylodynamic analysis of the Guangdong 2025 sequences and related ECSA-MAL genomes (dataset 3) was performed with BEAST X v10.5.0 [19]. A local molecular clock model was implemented to estimate separate background (2007–2024) and epidemic-period (2025) substitution rates. As the coalescent tree prior, a Bayesian SkyGrid was used for its smoother representation of effective population size (Ne) dynamics and improved computational efficiency, specified with 30 grid points; the number of grid points was selected to match the number of sequences (n = 247) and the time span of the dataset (2007–2025). Three independent MCMC chains of 100 million steps each were run, sampling every 50,000 steps; convergence was assessed in Tracer v1.7.2, and the maximum clade credibility tree was summarized with TreeAnnotator. The TMRCA for the Guangdong epidemic was calculated from the posterior distribution of the MCC tree, reported as mean, median, and 95% highest posterior density (HPD) intervals. Molecular clock rates were estimated separately for background and epidemic periods, reported as substitutions per site per year with 95% HPD intervals.

### 2.4 Lineage Fitness Analysis

Epidemic success and adaptive mutations were assessed using the Phylowave method [20], which computes a time-resolved index of epidemic success for each node based on pairwise genetic distances weighted by a geometric kernel with timescale parameter tau. The kernel timescale tau was set as 2.0 years given the best agreement with the lineage-level structure of the phylogeny; the 1-year time window matches surveillance resolution. Because deep divergence between CHIKV lineages can generate spurious index signals from neutrally accumulated substitutions, Phylowave was applied at two levels. First, as an unstratified baseline, a single analysis was run on the complete 2,410-genome time-scaled phylogeny. Second, the phylogeny was partitioned into its four major monophyletic lineages—the Asian urban lineage (AUL, n = 503), ECSA–Indian Ocean lineage (ECSA-IOL, n = 662), ECSA-MAL (n = 276), and ECSA– South American lineage (ECSA-SAL, n = 969)—and the minimal subtree of each lineage was analyzed independently with identical parameters (tau = 2.0 years, 1-year time window, minimum descendant count 50 and minimum group size 25, relaxed to 30 and 15 respectively for the smaller ECSA-MAL lineage; maximum 6 sublineages per lineage, 5 for ECSA-MAL). Following the Phylowave framework [20], sublineages with distinct index dynamics were detected by GAM-based segmentation with expanding time windows, using deviance explained and AIC as splitting criteria. A mutation was classified as lineage-defining when it was present in ≥80% of the genomes of a detected lineage and absent from the reconstructed ancestral amino acid state. The mutation lists from the two levels were then compared. The per-lineage results, which eliminate comparisons across the deep AUL–ECSA split, constituted the primary analysis. Lineage assignments were integrated into the full tree for visualization; lineage-defining mutations were mapped onto the CHIKV reference genome (strain S27), and the peak Phylowave index of each lineage was recorded as a measure of maximum epidemic success.

## Data Availability

All genome sequences have been deposited in the GenBase (https://ngdc.cncb.ac.cn/genbase), under accession number C_AA280545.1 to C_AA280583.1. Both sequence datasets and tree files (Auspice JSON) generated in this study have been deposited on GitHub (https://github.com/JingLu-GDIPH/CHIKV_Phy-Paper_2026/).

## 3 Results

### 3.1 Epidemiological Characteristics of the 2025 Guangdong Outbreak

Between July 8 (first laboratory-confirmed case) and December 21, 2025, a total of 23,464 CHIKV infections were reported across all 21 prefecture-level cities in Guangdong Province (Fig. 1a, https://cdcp.gd.gov.cn/ywdt/zdzt/yfjkkyr/yqxx/). The epidemic exhibited a distinctive biphasic temporal pattern. The first wave peaked during the week of July 20–26 at 2,940 reported cases per week, predominantly concentrated in Foshan municipality (Shunde district), where the first case of the outbreak were reported in Beijiao town. After a five-week decline reaching a trough of 178 cases per week (August 31–September 6), a second, larger wave emerged, peaking at 3,181 cases per week during September 28–October 4, driven by sustained transmission in Pengjiang and Heshan districts of Jiangmen city. The second wave accounted for 55.8% of all reported cases (13,103/23,464). Following the second peak, cases declined steadily over nine weeks, reaching zero by the week of December 21–27. All cases were clinically mild; no deaths were reported throughout the epidemic. Geographically, the outbreak was heavily concentrated in Foshan and Jiangmen, accounting for 8,980 (38.3%) and 10,035 (42.8%) of cumulative cases, respectively (Fig. 1b).

**Fig. 1.**
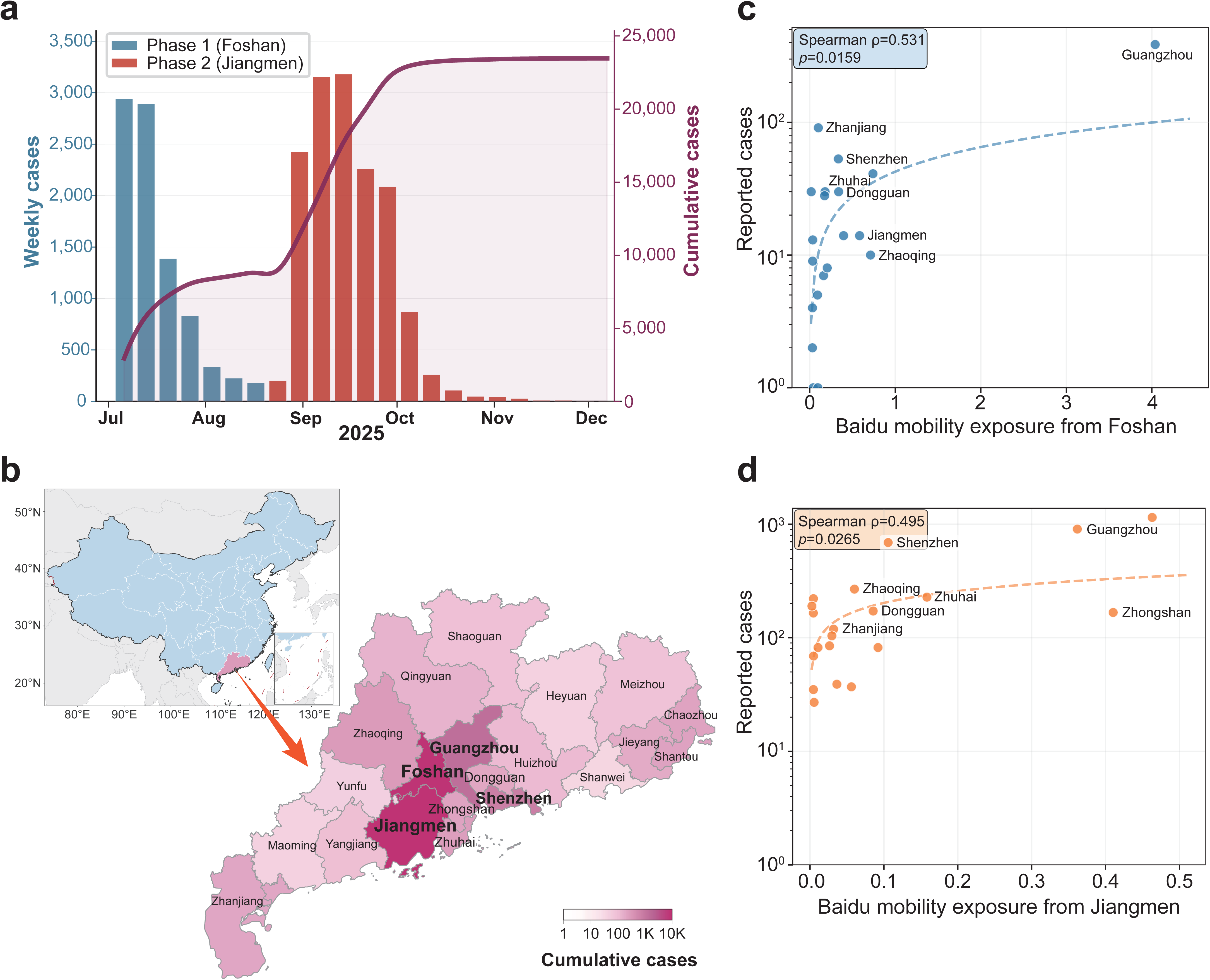
Epidemiological Characteristics of the 2025 Guangdong CHIKV Outbreak. Weekly epidemic curve. (a) and geographic distribution (b) of cumulative CHIKV cases across Guangdong Province’s 21 prefecture-level cities (July 8–December 21, 2025). Cities are colored by case count. The epicenter cities (Foshan and Jiangmen) are highlighted. Population Baidu mobility exposure explain the phase 1 (c) and phase 2 (d) spatial diffusion of reported CHIKV cases in Guangdong, 2025. City-level cases increased with Baidu mobility exposure from Foshan and from Jiangmen. Dashed lines show fitted log-log regression relationships.

Analysis of these mobility data showed that cities receiving higher population outflow from the phase-specific epicenter had significantly more reported CHIKV cases in both epidemic phases. Mobility exposure was positively associated with city-level case burden in Phase 1 (July–August, Foshan epicenter; Spearman *ρ* = 0.531, *p* = 0.0159) and Phase 2 (September–December, Jiangmen epicenter; Spearman *ρ* = 0.495, *p* = 0.0265) across 20 non-center prefecture-level cities in each phase (Fig. 1c-d). Mobility thus emerged as one of major correlates of CHIKV spatial spread.

### 3.2 Phylogenetic and Phylodynamic Analysis

To characterize the evolutionary dynamics and geographic distribution of CHIKV over the past two decades, we constructed a time-scaled phylogeny using 2,410 global genomes sampled during 2005 and 2026 from all endemic regions (Fig. 2a). Analysis of genotype distribution revealed dramatic shifts in CHIKV population structure. The West African (WA) genotype has become virtually extinct in public databases (2005–2026), likely due to sampling bias toward urban epidemics and limited surveillance in African forest regions.

**Fig. 2.**
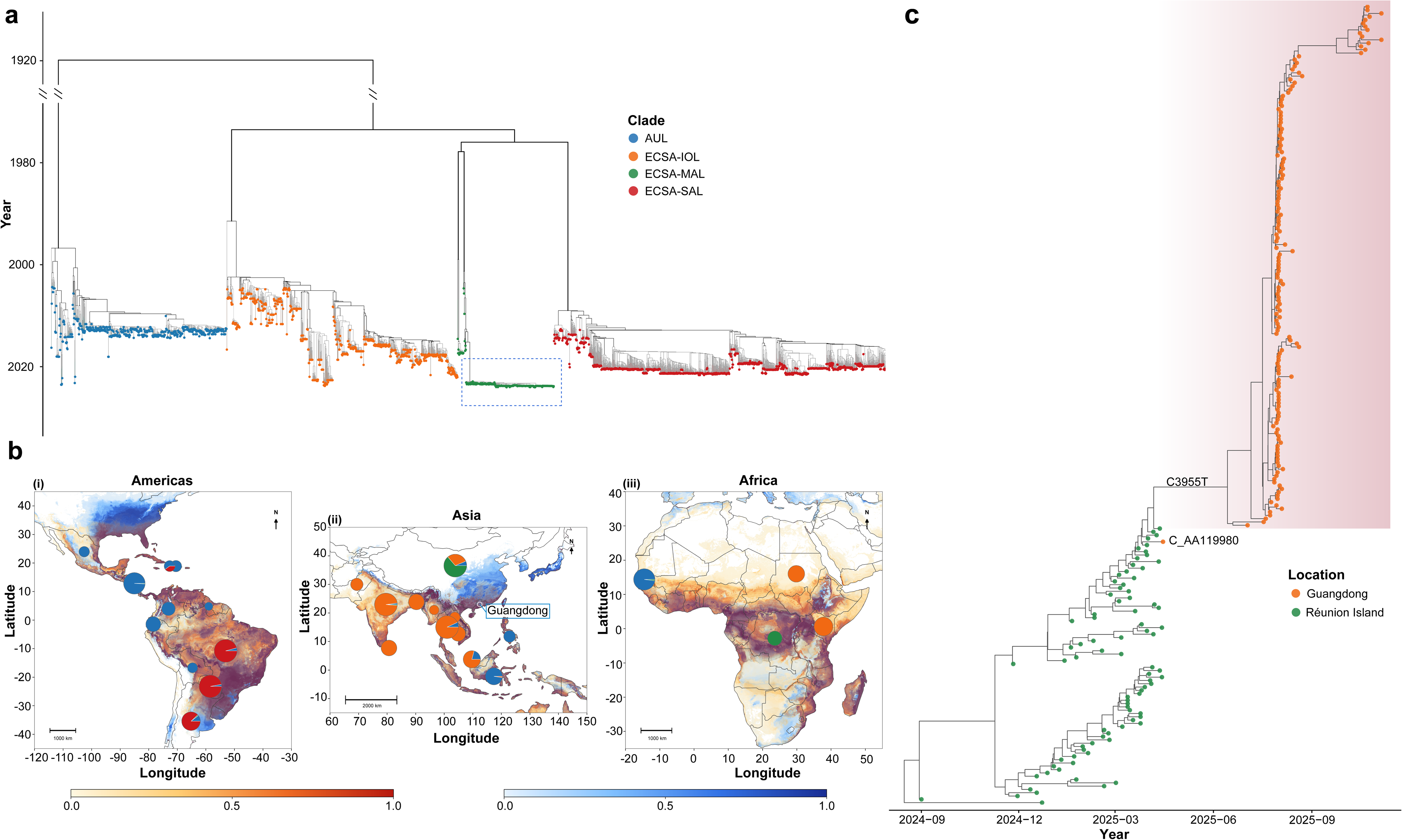
Phylogenetic Analysis of the 2025 Guangdong CHIKV Outbreak in Global Context and Geographic Distribution of CHIKV lineages. (a) Global CHIKV time-scaled phylogeny showing four major lineages: ECSA-SAL (South American lineage, red), ECSA-IOL (Indian Ocean Lineage, orange), ECSA-MAL (Middle African lineage, green), and AUL (Asian Urban lineage, blue). Sequences are shown as dots colored by lineage; the Guangdong 2025 outbreak sequences and related ECSA-MAL sequences, shown enlarged in panel (c), are highlighted by a rectangle. (b) Geographic distribution of CHIKV lineages across vector-defined ecological regions. Maps show the spatial distribution of major CHIKV lineages in relation to predicted ecological suitability for *Ae. aegypti* and *Ae. albopictus* across the Americas (i), Asia (ii), and Africa (iii) [28]. Background raster layers indicate vector occurrence probability, with warm colors representing *Ae. aegypti* suitability and blue colors representing *Ae. albopictus* suitability. Pie charts indicate the relative composition of CHIKV lineages among sampled genomes at each geographic location, and circle size reflects the number of available genomes from that location. Lineages are colored same as panel a. (c) Maximum clade credibility (MCC) tree of the Guangdong 2025 outbreak sequences (orange dots, rectangle-highlighted) combined with other related ECSA-MAL genomes (green dots), corresponding to the highlighted box in panel (a); the full tree is shown in Supplementary Figure S1.

The East/Central/South African (ECSA) genotype has undergone extensive diversification, differentiating into three distinct lineages whose geographic distributions, considered alongside vector-suitability patterns, reveal a clear ecological structure in CHIKV evolution (Fig. 2b): (1) ECSA-SAL (40.2% of sequences), dominant in *Ae. aegypti*-suitable regions of the Americas (Brazil, Paraguay, and Argentina) since its 2013–2014 invasion, where it has caused the American epidemics; (2) ECSA-IOL (27.5%), circulating across South and Southeast Asia, where it caused the 2004–2013 Indian Ocean and Indian subcontinent epidemics; and (3) ECSA-MAL (11.5%), concentrated in central Africa and East Asia—regions of comparatively high *Ae. albopictus* suitability—and associated with sporadic central African activity, the 2024–2025 Indian Ocean resurgence [8], and the 2025 Guangdong outbreak in mainland China. The AUL lineage (20.9% of sequences) has caused recurrent outbreaks in Southeast Asia since the 1950s, including the 2018–2019 re-emergence, and—together with ECSA-SAL—predominates in *Ae. aegypti*-suitable regions of the Americas; these parallel histories underscore that lineage persistence may not be necessarily reflected by the number of available genomes. Within this ecological framework, the Guangdong outbreak—an *Ae. albopictus*-driven epidemic in a subtropical East Asian setting—is consistent with the broader lineage-vector pattern and reinforces the need for region-specific monitoring of lineage–vector combinations.

The 2025 Guangdong outbreak sequences belong to the ECSA-MAL Lineage and form a star-like phylogeny, strongly supporting a single introduction event followed by rapid epidemic expansion (Fig. 2c, full tree presented in Supplementary Figure S1). Within the global phylogeny, the Guangdong outbreak cluster is defined by a single synonymous substitution (C3955T) relative to its closest relatives from French Indian Ocean territories (Réunion island) sampled in early 2025—most parsimoniously a neutral founder mutation, or lineage “tag”, present in (or arising immediately before) the genome that seeded the outbreak [8]. This lineage has been circulating since at least 2007 but sampling has been sparse, with only 106 sequences available in public databases prior to the 2025 Guangdong outbreak, the majority of which are from French Indian Ocean territories. This sampling gap underscores substantial surveillance deficiencies in regions where CHIKV is endemic.

Bayesian Skygrid analysis suggested an increase in the effective population size of the CHIKV MAL lineage around 2017, followed by a renewed expansion during the 2024–2025 epidemic period (Fig. 3a). However, the wide 95% HPD intervals, particularly across the post-2017 period, indicate substantial uncertainty, likely driven by sparse genomic sampling and a molecular surveillance gap. Therefore, although the analysis supports recent epidemic expansion, the detailed evolutionary trajectory between the earlier expansion and the 2025 outbreak remain incompletely resolved. Because over half of the genomes were sampled within a single year, a local molecular clock was used (see Methods); stepping-stone sampling showed decisive support for it over a strict clock (log marginal likelihood −22,644.5 vs −23,335.3; ln BF = 690.8). The epidemic-period substitution rate showed a 7.1-fold acceleration relative to the historical background. The background clock rate was estimated at 5.95×10^-4^ substitutions/site/year (95% HPD: 5.17–6.73×10^-4^), whereas the outbreak-specific rate escalated to 4.21×10^-3^ substitutions/site/year (95% HPD: 3.54–4.91×10^-3^) (Fig. 3b). This transient acceleration is consistent with time-dependent rate estimation during rapid, densely sampled transmission (see Discussion).

**Fig. 3.**
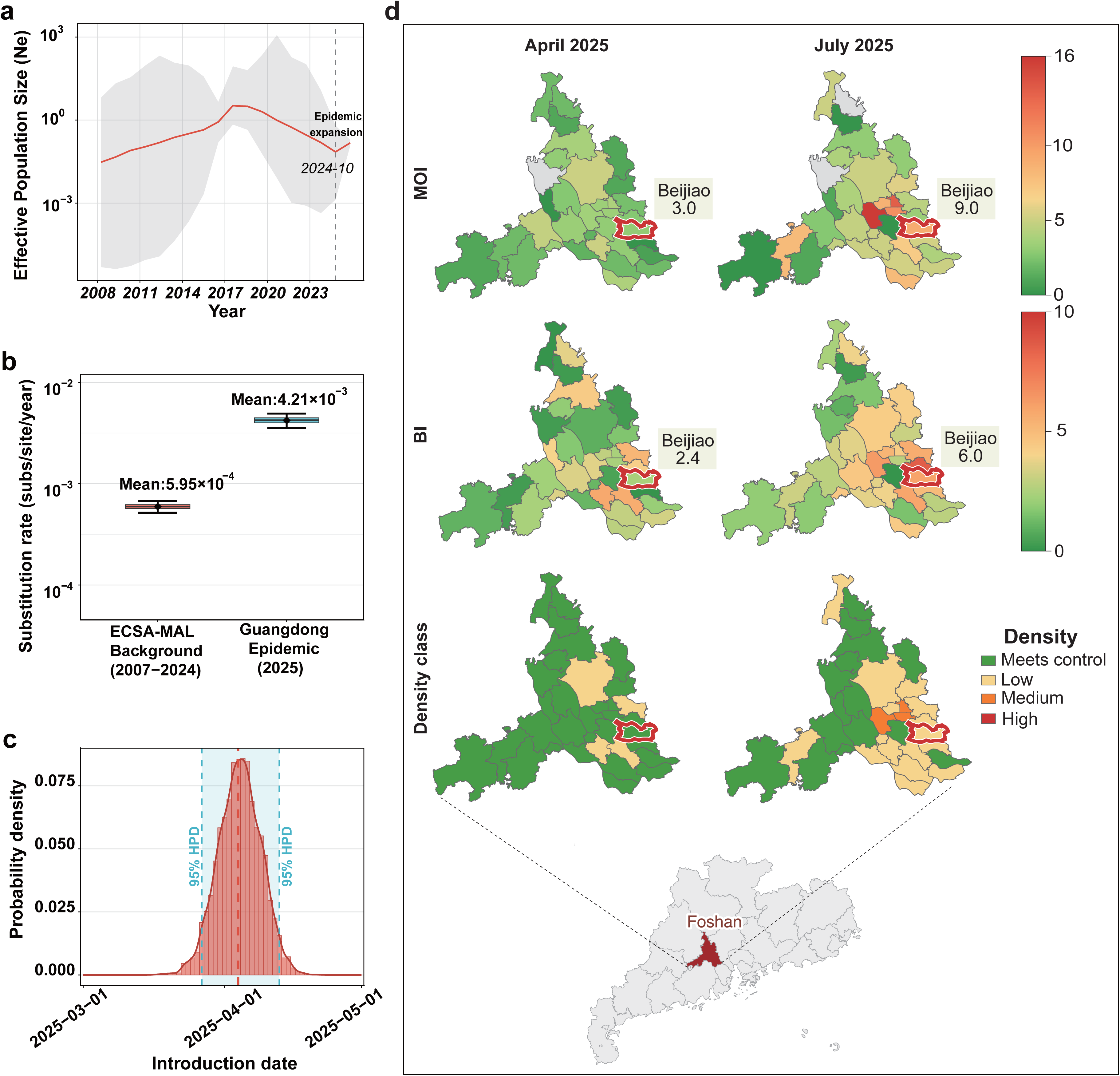
Phylodynamic Analysis of Guangdong related ECSA-MAL dataset (see Methods for detail) and Vector Density Correlates. (a) Bayesian Skygrid showing effective population size (Ne) changes through time for the Guangdong related ECSA-MAL clade (2007–2025). The median estimate (solid red line) and 95% highest posterior density (HPD) intervals (shaded area) are shown. (b) Bar chart comparing evolutionary rates of Guangdong 2025 sequences and related ECSA-MAL clade genomes estimated using a local clock model in BEAST, allowing separate rate estimates for the background period (2007–2024) and the epidemic period (2025). The background rate (red bar) was estimated at 5.95×10^-4^ substitutions/site/year (95% HPD: 5.17–6.73×10^-4^), while the epidemic rate (blue bar) was at 4.21×10^-3^ substitutions/site/year (95% HPD: 3.54–4.91×10^-3^). Error bars represent 95% HPD intervals. (c) Introduction time estimation density curve. Smoothed kernel density estimate of the TMRCA posterior distribution with 95% HPD interval (blue shaded area, March 26–April 12, 2025). The median introduction date (April 5, 2025) is marked with a vertical red line. (d) *Ae. albopictus* density dynamics in Foshan city of Guangdong Province during both April and June 2025, shown by surveillance-based Mosquito Ovitrap Index (MOI) and Breteau Index (BI) indices and aligned with epidemic transmission phases. Beijiao town—where the first case was reported and from which the outbreak originated—is highlighted with a red border. MOI represents the number of positive mosquito ovitraps per 100 valid recovered ovitraps, and BI represents the number of positive water containers per 100 households surveyed. According to standard vector risk assessment criteria, a BI/MOI < 5 meets the threshold for effective epidemic control, while 5 ≤ BI/MOI < 10 indicates low vector density, 10 ≤ BI/MOI < 20 indicates medium density, and BI/MOI ≥ 20 indicates high density.

According to the inferred time of the most recent common ancestor (TMRCA) for the epidemic, the estimated median introduction date was early April 2025 (April 5, 2025), with the 95% HPD interval spanning from March 26 to April 12, 2025 (Fig. 3c). The inferred introduction window coincides with the plateau phase of the concurrent Réunion Island epidemic [8], and the Guangdong outbreak cluster shares its closest ancestry with ECSA-MAL viruses circulating in the French Indian Ocean territories, consistent with importation from this region during a period of intense epidemic activity. Notably, a CHIKV sequence (C_AA119980, GenBase) sampled on April 15, 2025 by Guangzhou Customs, from an imported case in Guangdong, clusters within ECSA-MAL but on a sister branch, differing from the outbreak cluster by five nucleotide substitutions. Given that this imported case falls very close to the inferred introduction window (95% HPD: March 26–April 12, 2025), it may represent a co-introduction event from the same source population. While the C_AA119980-associated case was identified through symptomatic presentation, the outbreak-founding strain appeared to have established cryptic transmission, possibly entering through an asymptomatic or subclinical infection that evaded early detection and subsequently seeded sustained local transmission.

The first CHIKV case was reported on June 22, 2025, approximately 2.5 months post the inferred introduction date. This discrepancy suggests a period of undetected low-level transmission spanning April through mid-June 2025. The *Ae. albopictus* surveillance data provide a temporal pattern that possibly explain the surveillance gap (Fig. 3d). In the outbreak town of Beijiao (Shunde, Foshan), the MOI increased from 3.0 in April to 9.0 in June and remained at this level in July, close to the medium-density threshold (detailed in Supplementary Table S2). These patterns suggest that CHIKV may have been introduced during April–June, when vector density remained relatively low and early transmission could have gone undetected. The seasonal rise in *Aedes* density during June–July likely created more favorable conditions for amplification, contributing to the detectable outbreak in Beijiao.

### 3.3 Per-lineage Phylowave Analysis Identifies Candidate Adaptive Mutations

Phylowave uses phylogenetic structure and sampling time to estimate a sublineage-specific index that reflects the relative contribution of a lineage to future viral diversity [20]. To identify shifts in molecular epidemiological dynamics within major CHIKV lineages, we applied a whole-tree and a per-lineage Phylowave analysis. The unstratified whole-tree analysis identified 133 candidate lineage-defining mutations but resolved only four coarsely defined lineages. Application of the per-lineage Phylowave identified 14 epidemic sublineages with rising index across the four major lineages (AUL 3, ECSA-IOL 5, ECSA-MAL 4, ECSA-SAL 2) (Supplementary Figure S2). The rising index values indicate sublineages that have gained evolutionary or epidemiological prominence—through enhanced transmissibility, ecological opportunity, founder effects, or adaptive changes—and should be interpreted as candidate expansion-associated lineages rather than direct evidence of beneficial mutations. To refine this inference, we subsequently identified lineage-defining mutations and examined whether these changes were enriched in high-index sublineages, thereby linking shifts in lineage dynamics to putative adaptive variation.

The per-lineage analysis yielded 33 lineage-defining mutations across nine of the ten CHIKV proteins (Supplementary Table S3 and Supplementary Figure S2). The 33 mutations included all six mutations with previous literature support (including the four experimentally validated substitutions E1:A226V, E1:K211E, E2:V264A, and E1:A98T) and 12 substitutions marking the deeply divergent AUL lineage—while 115 of the 133 whole-tree candidates were eliminated as artifacts of cross-lineage divergence. Conversely, 15 mutations were detected exclusively by the per-lineage analysis (13 in ECSA-IOL lineages L2–L4 and 2 in ECSA-SAL L1, Supplementary Table S3), none of which has been previously described. These signals were masked at the whole-tree level because the corresponding sublineages were not resolved there; for example, the nine defining mutations of the expanding ECSA-IOL L4 lineage are present in 93–100% of L4 genomes but reached only ≈14% within the whole-tree IOL lineage into which L4 was absorbed, far below the 80% threshold.

In per-lineage phylowave analysis, several well-characterized CHIKV adaptive mutations were recovered with high peak Phylowave index values (Fig. 4a). The E1:A226V substitution, which confers enhanced replication in *Ae. albopictus* [21], was identified in ECSA-IOL lineage L2 with a peak index of 0.831—the highest among all lineages. The compensatory mutations E1:K211E and E2:V264A [22], which emerged following A226V acquisition, were identified in ECSA-IOL L3 with a peak index of 0.797. The E1:A98T substitution, subject to epistatic constraints in Asian lineage viruses [23], was detected in ECSA-SAL L1 with a peak index of 0.681. Mutations were distributed across NSP3 (7/33, 21.2%, concentrated in the hypervariable domain), envelope proteins E2 (7) and E1 (6), NSP4 (5), NSP2 (3), NSP1 (3), and structural proteins E3 and 6K (one each). Notably, 12 mutations were exclusive to the AUL L1 lineage, reflecting the independent evolutionary trajectory of the Asian genotype.

**Fig. 4.**
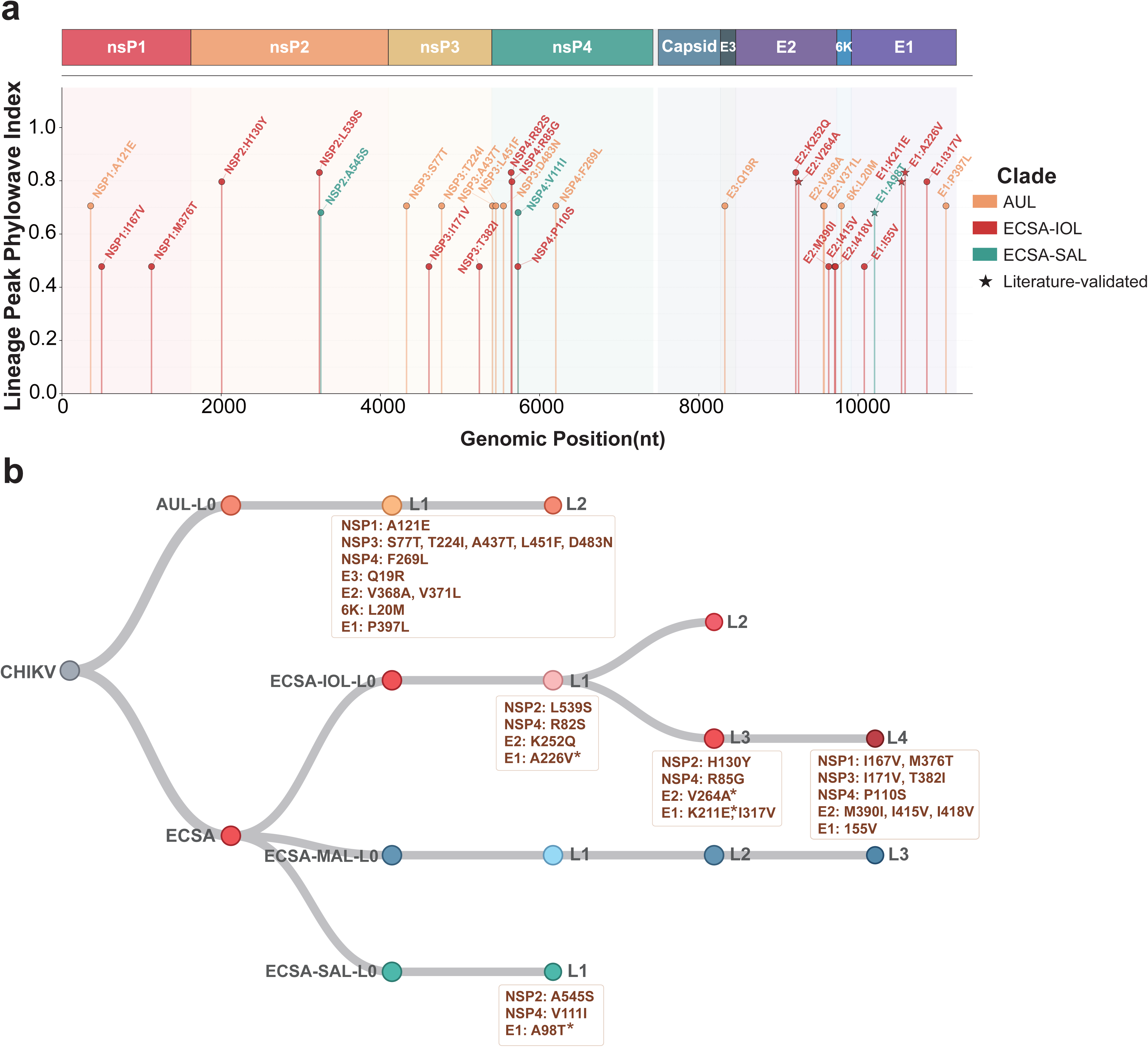
Genomic Architecture of CHIKV Adaptive Mutations Identified by Per-Lineage Phylowave Analysis. (a) Lollipop plot of 33 clade-defining mutations resolved by the per-lineage analysis displayed along the CHIKV genome (NSP1–4, Capsid, E3, E2, 6K, E1). Gene boxes are colored by protein. The y-axis shows the peak Phylowave index of the corresponding lineage branch, representing maximum epidemic success. Mutation markers are colored by lineage: AUL (orange), ECSA-IOL (red), ECSA-SAL (teal). (b) Phylowave Lineage Schema and Candidate adaptive Mutations in CHIKV Evolution. Phylowave Index analysis showing evolutionary relationships among 2,410 sequences across four major lineages. Nodes are colored by lineage. Candidate adaptive mutations are displayed on connecting branches, grouped by gene. Star markers (*) indicate literature-validated adaptive mutations (E1:A226V, E1:K211E, E2:V264A, E1:A98T).

Several lineage-defining mutations mark recurrent functional hotspots. Phylogenetic analysis revealed that NSP3:D483N in the AUL lineage and NSP3:N483S in the ECSA-MAL lineage represent independent substitutions at the same hotspot position in the hypervariable domain adjacent to the FGDF-1 G3BP-binding motif. Similarly, NSP3:T224I occurred independently in both AUL and ECSA-SAL lineages within the zinc finger domain, suggesting convergent evolution toward optimal replication complex function. E2:V371L, situated within the E2 transmembrane anchor, probably represent AUL-specific adaptations. In ECSA-SAL, NSP4:V111I (peak index 0.681) at the RdRp N-terminal/domain boundary and NSP2:A545S (peak index 0.681) in the protease domain may modulate polymerase conformation and replicase assembly, respectively. The ECSA-MAL lineage produced the highest fitness signal (peak index 0.999) in 2024–2025. However, interpretation of sublineage structure is complicated by significant surveillance gaps (2012–2015, 2020–2023). No lineage-defining mutations were detected, consistent with either high genetic homogeneity or insufficient sampling to capture evolutionary transitions (tau=2 years). The Guangdong outbreak thus appears driven by epidemiological factors (vector abundance, host susceptibility) rather than detectable viral genetic innovation, though limited historical sampling precludes definitive conclusions about adaptive evolution.

## 4 Discussion

This study provides the genomic and epidemiological characterization of the largest CHIKV outbreak in mainland China, contextualizing it within global CHIKV lineages evolution patterns. Our integrated analysis has three key findings. First, phylodynamic analysis of the ECSA-MAL lineage revealed approximately 2.5 months of cryptic transmission before the first case detection, enabled by sub-threshold vector densities during the spring introduction period. Second, population mobility from phase-specific epicenters was moderately associated with city-level case burden in both phases, indicating that human movement contributed to spatial diffusion but was constrained by vector ecology and local transmission conditions. Third, global phylogenetic analysis uncovered substantial surveillance gaps in the ECSA-MAL lineage and revealed potential distinct regional adaptive strategies underscoring the need for ecologically informed molecular surveillance.

The long internal branch of the ECSA-MAL lineage (2007–2024), with a few sequences available in public databases prior to the Guangdong outbreak, reveals critical surveillance deficiencies for this lineage in endemic regions. Notably, CHIKV is not a mandatory notifiable disease in most countries, resulting in substantial underreporting, particularly the clinical diagnosis is complicated by syndromic overlap with dengue and Zika virus infections [24]. The ECSA-MAL lineage has been poorly sampled despite its endemic circulation in French Indian Ocean territories (Réunion, Mayotte), where a resurgence was documented during 2024–2025 [8]. The associated epidemiological data were insufficient to risk assess for countries with importation risk. This disconnect between genomic data generation and epidemiological contextualization represents a missed opportunity for endemic outbreak preparedness. The 2.5-month cryptic transmission period in Guangdong demonstrates how arboviruses can establish low-level transmission chains below the detection threshold when vector densities are sub-optimal. Meta-analytic evidence indicates that approximately 25% of CHIKV infections are asymptomatic [25], enabling subclinical transmission that evades symptom-based surveillance systems.

Population mobility from the two phase-specific epicenters, Foshan and Jiangmen, was moderately associated with CHIKV case distribution, but it did not provide a deterministic explanation of spatial spread (Fig.1c-d). This contrasts with respiratory viruses such as SARS-CoV-2, for which population flow can account for much larger spatial variation. For example, Jia et al. [26] reported that population outflow from Wuhan during the early COVID-19 epidemic was strongly correlated with confirmed cases across 296 Chinese prefectures (Pearson r = 0.952; R^2^ = 0.906), indicating that human mobility explained over 90% of spatial variation in SARS-CoV-2 case distribution. This discrepancy arises from ecological factors unique to arbovirus transmission. The limited dispersal range of *Ae. albopictus* (100–500 m) [27] means that local transmission is geographically constrained around mosquito breeding sites, regardless of where infected humans travel. The biphasic epidemic pattern observed in Foshan and Jiangmen illustrates the spatial dynamics of arbovirus spread: human mobility seeds new locations, while local vector ecology determines whether sustained transmission is established. Outside Guangdong, spillover to other provinces remained limited. Neighboring provinces—including Guangxi, Fujian, and Hunan—reported only sporadic cases or small clusters, most of which were epidemiologically linked to travel from Guangdong. The limited geographic expansion of CHIKV in mainland China highlights that virus transmission fundamentally requires competent vector populations; human mobility can only initiate outbreaks where local vector abundance and ecology permit. For importation-risk countries such as China, three practical measures are suggested. First, because local transmission is possible only during the vector-active season (approximately April/May through November in Guangdong), post-arrival interventions should be prioritized within this window: travelers arriving from epidemic or endemic regions should receive symptom-awareness guidance and be offered diagnostic testing within 1–2 weeks of arrival. Second, and most importantly, returning travelers at risk should be explicitly advised to rigorously avoid mosquito bites during the first weeks after arrival—by wearing long-sleeved clothing, applying topical repellents, and sleeping under bed nets—so that viremic travelers (including asymptomatic ones) do not infect local vectors. Third, pre-season arbovirus surveillance during spring months, when vector densities are below the transmission threshold, is critical for early detection of viral introductions before summer amplification.

The lineage-specific molecular epidemiology and geographic distribution highlights the need for lineage specificity of candidate adaptive mutations analysis. The 33 candidate mutations were identified from the global dataset of 2,410 genomes via per-lineage Phylowave analysis. The convergence of Phylowave-detectable epidemic success with experimentally validated adaptive mutations supports the biological relevance of this approach. The mutations associated with the highest peak index including E1:A226V and E1:K211E/E2:V264A, enabled efficient *Ae.Albopictus* transmission or restored fitness in *Ae. aegypti* following A226V acquisition [21,22]. The 12 novel lineage-defining mutations identified in AUL L1 (Fig. 4b), particularly NSP3:T224I and E2:V371L, warrant experimental validation through reverse genetics and vector competence studies. The rising index trajectory of AUL L1 suggests continued expansion, consistent with the re-emergence of CHIKV in Southeast Asia during 2018–2019. *Ae.aegypti* dominated regions should monitor for ECSA-SAL characteristic mutations (particularly in the NSP3 hypervariable domain), while *Ae. albopictus* dominated regions should track E1:A226V-containing lineages and their derivative adaptations. The candidate adaptive mutations identified here currently represent computational predictions derived from the phylogenetic association between mutations and lineage epidemic success. Definitive demonstration of their functional roles will require reverse-genetics reconstruction of mutant viruses, replication-kinetics assays in mammalian and mosquito cells, vector-competence experiments in *Ae. aegypti* and *Ae. albopictus*, and, where warranted, animal models.

As CHIKV continues to expand its geographic range in the context of climate change, urbanization, and increasing human mobility, the lessons from this outbreak are globally relevant. Enhanced genomic surveillance integrated with epidemiological and entomological data—particularly during pre-peak months—will be critical for early detection and rapid response. Future work should focus on functional validation of candidate adaptive mutations, dissection of epistatic interactions across vector species, and development of predictive models linking viral genotypes to outbreak risk in different ecological settings.

## Acknowledgments

This work was supported by National Major Project of the “Surveillance and Early Warning of Emerging and Acute Infectious Diseases” Initiative (Grant No. 2026ZD01909500), General Program of the National Natural Science Foundation of China (Grant No. 82574220) and Guangdong Provincial Center for Disease Control and Prevention Supports Talent Projects (Grant No. 0720240122).

## Author contributions

L.Y. and J.L. contributed to the conception of the study. S.X., X.H., J.H., M.C., H.L., Y.H., C.Z., G.Z., S.T. and S.G. performed the experiments and the data analyses. J.L., S.X. and Z.L. performed bioinformatic analysis. L.Y., S.X., and J.L. wrote the manuscript. All authors have read and approved the final manuscript.

## References

1. Weaver SC, Lecuit M. Chikungunya virus and the global spread of a mosquito-borne disease. N Engl J Med 2015;372(13):1231–9. 10.1056/NEJMra1406035.

2. Lim A, Shearer FM, Sewalk K et al. The overlapping global distribution of dengue, chikungunya, zika and yellow fever. Nat Commun 2025;16(1):3418. 10.1038/s41467-025-58609-5.

3. Ning X, Xia B, Wang J et al. Host-adaptive mutations in chikungunya virus genome. Virulence 2024;15(1):2401985. 10.1080/21505594.2024.2401985.

4. Spicher T, Delitz M, Schneider A de B, et al. Dynamic molecular epidemiology reveals lineage-associated single-nucleotide variants that alter RNA structure in chikungunya virus. Genes 2021;12(2):239–55. 10.3390/genes12020239.

5. Luo L, Liang H ying, Hu Y shan, et al. Epidemiological, virological, and entomological characteristics of dengue from 1978 to 2009 in Guangzhou, china. J Vector Ecol 2012;37(1):230–40. 10.1111/j.1948-7134.2012.00221.x.

6. Liu Y, Wang X, Tang S et al. The relative importance of key meteorological factors affecting numbers of mosquito vectors of dengue fever. PLoS Negl Trop Dis 2023;17(4):e0011247. 10.1371/journal.pntd.0011247.

7. World Health Organization. Chikungunya virus disease-Global situation. 3 Oct. 2025. https://www.who.int/emergencies/disease-outbreak-news/item/2025-DON581.

8. Frumence E, Piorkowski G, Traversier N et al. Genomic insights into the re-emergence of chikungunya virus on Réunion Island, France, 2024 to 2025. Eurosurveillance 2025;30(22):2500344. 10.2807/1560-7917.ES.2025.30.22.2500344.

9. D W, J W, Q Z, et al. Chikungunya outbreak in guangdong province, china, 2010. Emerg Infect Dis 2012;18(3):1–3. 10.3201/eid1803.110034.

10. Pan J, Fang C, Yan J et al. Chikungunya fever outbreak, zhejiang province, china, 2017. Emerg Infect Dis 2019;25(8):1589–91. 10.3201/eid2508.181212.

11. Liu H, Zheng W, Holmes EC et al. Spatiotemporal dynamics of the unprecedented chikungunya outbreak in Guangdong, China. Emerg Microbes Infect 2026;15(1):2695536. 10.1080/22221751.2026.2695536.

12. He Y, Zeng C, Tan S et al. Viral kinetics of the chikungunya virus ESCA-MAL lineage during the 2025 guangdong outbreak. Virol Sin 9 June 2026:S1995–820X(26)00093-3. 10.1016/j.virs.2026.06.004.

13. Lu J, Plessis L du, Liu Z, et al. Genomic Epidemiology of SARS-CoV-2 in Guangdong Province, China. Cell Apr. 2020:S0092867420304864. 10.1016/j.cell.2020.04.023.

14. Li H. Minimap2: pairwise alignment for nucleotide sequences. Bioinformatics 2018;34(18):3094–100. 10.1093/bioinformatics/bty191.

15. Li H. A statistical framework for SNP calling, mutation discovery, association mapping and population genetical parameter estimation from sequencing data. Bioinformatics 2011;27(21):2987–93. 10.1093/bioinformatics/btr509.

16. Katoh K, Standley DM. MAFFT multiple sequence alignment software version 7: improvements in performance and usability. Mol Biol Evol 2013;30(4):772–80. 10.1093/molbev/mst010.

17. Minh BQ, Schmidt HA, Chernomor O et al. IQ-TREE 2: New Models and Efficient Methods for Phylogenetic Inference in the Genomic Era. Mol Biol Evol 2020;37(5):1530–4. 10.1093/molbev/msaa015.

18. Sagulenko P, Puller V, Neher RA. TreeTime: maximum-likelihood phylodynamic analysis. Virus Evol 2018;4(1):vex042. 10.1093/ve/vex042.

19. Baele G, Ji X, Hassler GW et al. BEAST X for bayesian phylogenetic, phylogeographic and phylodynamic inference. Nat Methods 2025;22(8):1653–6. 10.1038/s41592-025-02751-x.

20. Lefrancq N, Duret L, Bouchez V et al. Learning the fitness dynamics of pathogens from phylogenies. Nature 2025;637(8046):683–90. 10.1038/s41586-024-08309-9.

21. Tsetsarkin KA, Vanlandingham DL, McGee CE et al. A single mutation in chikungunya virus affects vector specificity and epidemic potential. PLOS Pathog 2007;3(12):e201. 10.1371/journal.ppat.0030201.

22. Agarwal A, Sharma AK, Sukumaran D et al. Two novel epistatic mutations (E1:K211E and E2:V264A) in structural proteins of chikungunya virus enhance fitness in aedes aegypti. Virology 2016;497:59–68. 10.1016/j.virol.2016.06.025.

23. Tsetsarkin KA, Chen R, Leal G et al. Chikungunya virus emergence is constrained in Asia by lineage-specific adaptive landscapes. Proc Natl Acad Sci U S A 2011;108(19):7872–7. 10.1073/pnas.1018344108.

24. Waggoner JJ, Gresh L, Mohamed-Hadley A et al. Single-reaction multiplex reverse transcription PCR for detection of zika, chikungunya, and dengue viruses. Emerg Infect Dis 2016;22(7):1295–7. 10.3201/eid2207.160326.

25. Salazar Flórez JE, Restrepo BN, Freitas LP et al. Spatio-temporal analysis of the distribution and co-circulation of dengue, chikungunya, and zika in medellín, colombia, from 2013 to 2021. PLoS Negl Trop Dis 2025;19(9):e0013470. 10.1371/journal.pntd.0013470.

26. Jia JS, Lu X, Yuan Y et al. Population flow drives spatio-temporal distribution of COVID-19 in China. Nature 2020;582(7812):389–94. 10.1038/s41586-020-2284-y.

27. Sedda L, Vilela APP, Aguiar ERGR et al. The spatial and temporal scales of local dengue virus transmission in natural settings: a retrospective analysis. Parasit Vectors 2018;11:79–92. 10.1186/s13071-018-2662-6.

28. Kraemer MU, Sinka ME, Duda KA et al. The global distribution of the arbovirus vectors Aedes aegypti and Ae. albopictus. eLife 2015;4:e08347. 10.7554/eLife.08347.

