## Supplementary for "Genomic Epidemiology of CHIKV During the Largest Outbreak in Mainland China-Implication for CHIKV Intervention"

**Supplementary Figure S1** Extended maximum clade credibility tree of the Guangdong 2025 outbreak sequences and related ECSA-MAL genomes, expanding the tree shown in Fig. 2c. The tree was inferred from a 247-genome dataset comprising 159 Guangdong 2025 genomes, 83 Réunion Island 2024–2025 genomes, and earlier-diverging ECSA-MAL sister-branch sequences sampled in central Africa (Gabon, 2007; Cameroon, 2016–2018) and France (2017). This extended view places the Guangdong outbreak clade within the full temporal context of the ECSA-MAL lineage.

**Supplementary Figure S2** Time-scaled phylogeny of CHIKV genomes annotated by Phylowave-defined lineages. The time-scaled maximum clade credibility phylogeny shows 2,410 CHIKV genomes assigned to Phylowave-defined lineages across the four major clades: AUL, ECSA-IOL, ECSA-MAL, and ECSA-SAL. Tips are colored according to Phylowave lineage assignment. Lineages labeled L0 represent background or ancestral groups within each clade, whereas L1–L4 represent epidemic or expansion-associated lineages detected by the per-lineage Phylowave analysis.

**Supplementary Table S1** Sequencing and genome assembly metrics for CHIKV isolates from the 2025 Guangdong outbreak, China. Detailed per-sample quality and assembly parameters for viral isolates (Ct≤35) sequenced on the Illumina MiSeq platform. Reported metrics include RT-PCR cycle threshold (Ct) values, clean data volume (Gb), Q30 base percentage, mapping rate, mean sequencing depth, genome breadth of coverage (≥10×), assembled genome length, and percentage coverage.

**Supplementary Table S2** Township-level *Aedes albopictus* surveillance indices in Foshan City, April–July 2025. The Mosquito Ovitrap Index (MOI, positive ovitraps per 100 ovitraps deployed) and Breteau Index (BI, positive containers per 100 households surveyed) are shown by calendar month for all 32 towns/subdistricts of the five districts; the July 2025 CHIKV outbreak focus, Beijiao town in Shunde district, is flagged. “—” indicates no valid records for that month. Ovitraps deployed and households surveyed are totals for April–July.

**Supplementary Table S3** Catalog of 33 lineage-defining mutations identified by per-lineage Phylowave analysis, with functional-domain annotations, peak index values, frequency data (within the defining lineage and in the remaining genomes of the clade), enrichment scores (Inf, background frequency of zero), lineage assignments, and the level of analysis at which each mutation was detected—shared with the whole-tree analysis (n = 18) or detected exclusively by the per-clade analysis (n = 15).
