## Supplementary figures and images for "Genomic Epidemiology of CHIKV During the Largest Outbreak in Mainland China-Implication for CHIKV Intervention"

### Supplementary Figure S1

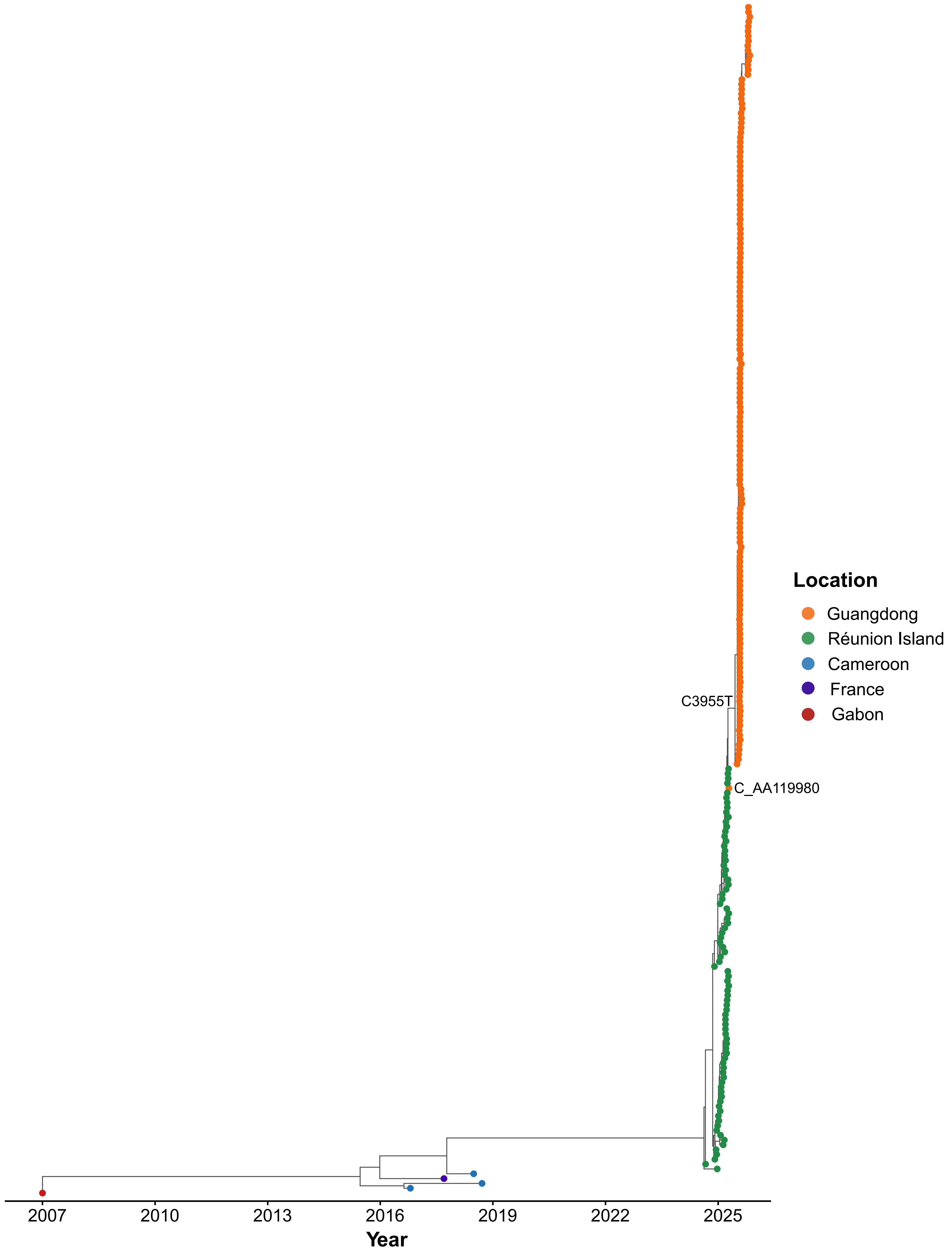

### Supplementary Figure S2

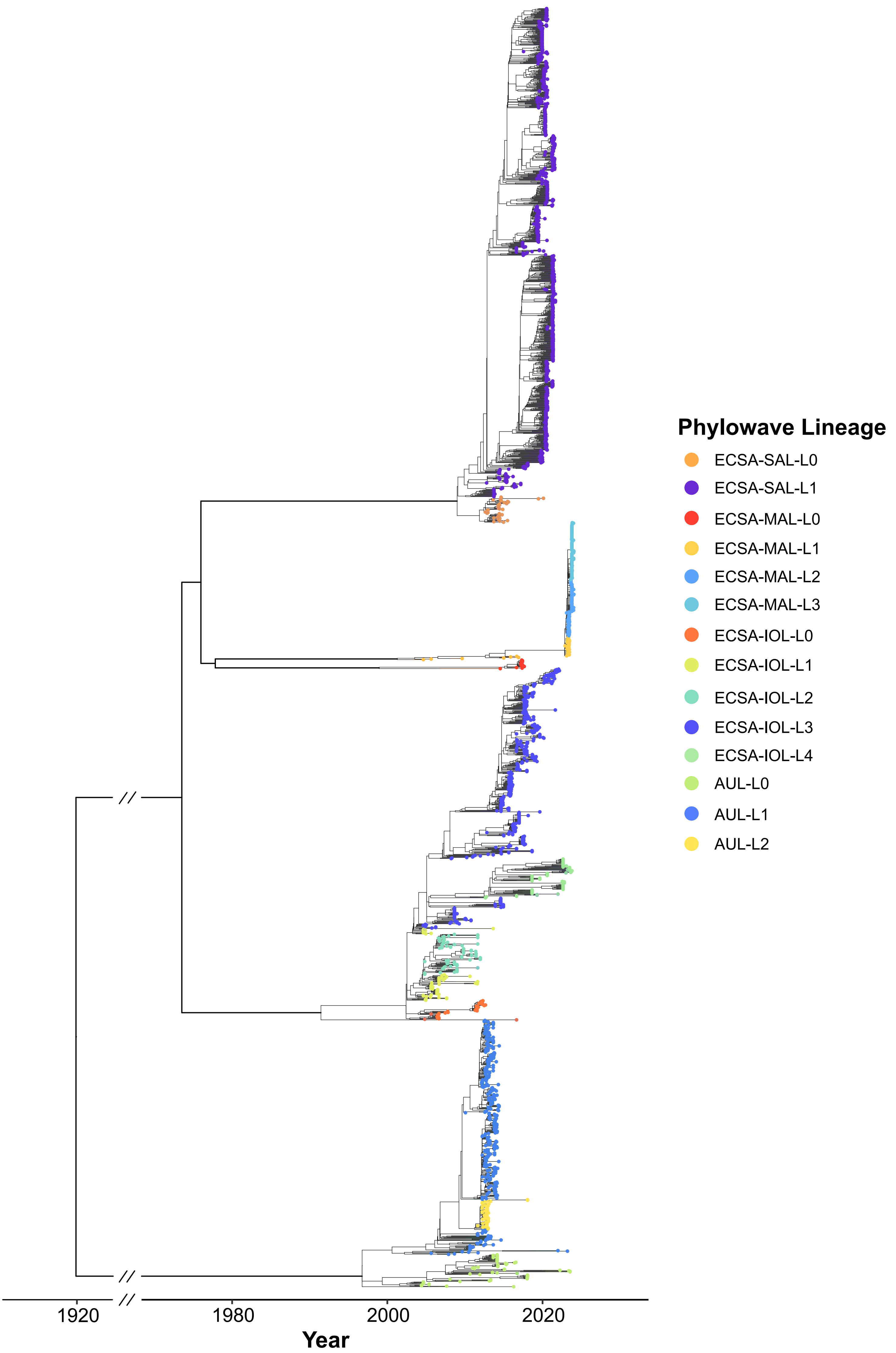
